# Trends in non-COVID-19 hospitalizations prior to and during the COVID-19 pandemic period, United States, 2017 – 2021

**DOI:** 10.1101/2022.04.26.22274301

**Authors:** Kelsie Cassell, Casey M Zipfel, Shweta Bansal, Daniel M. Weinberger

## Abstract

COVID-19 pandemic-related shifts in healthcare utilization, in combination with trends in non-COVID-19 disease transmission and NPI use, had clear impacts on infectious and chronic disease hospitalization rates. Using a national healthcare billing database (C19RDB), we estimated the monthly incidence rate ratio of hospitalizations between March 2020 and June 2021 according to 19 ICD-10 diagnostic chapters and 189 subchapters. The majority of hospitalization causes showed an immediate decline in incidence during March 2020. Hospitalizations for diagnoses such as reproductive neoplasms, hypertension, and diabetes returned to pre-pandemic norms in incidence during late 2020 and early 2021, while others, like those for infectious respiratory disease, never returned to pre-pandemic norms. These results are crucial for contextualizing future research, particularly time series analyses, utilizing surveillance and hospitalization data for non-COVID-19 disease. Our assessment of subchapter level primary hospitalization codes offers new insight into trends among less frequent causes of hospitalization during the COVID-19 pandemic.

## Introduction

The emergence of COVID-19 in the United States had immediate impacts on healthcare utilization and rates of disease due to non-SARS-CoV-2 related illness. Between March and April 2020, when a majority of US states had enacted stay-at-home orders, non-COVID-19 related illness dropped approximately 20-30% [1,2]. These shifts in hospitalization rates could be attributed to a combination of factors influencing health and healthcare. During the early months of the pandemic, an estimated 41% of US adults avoided medical care due to COVID-19 related concerns [3]. At the same time, there were massive and long-lasting alterations to daily activities and contact patterns. Non-essential workers were required to work-from-home, businesses closed, students transitioned to remote learning, and outpatient healthcare settings favored telehealth visits over in-person visits when possible [4,5]. These sudden shifts, in addition to pathogen-pathogen interference, and non-pharmaceutical interventions (NPI) like face masks, had the potential to influence the transmission of other pathogens and to both directly and indirectly affect the rates of hospitalization for a multitude of non-COVID-19 conditions [6,7].

The incidence of many infectious diseases decreased during the pandemic period (starting in March 2020) compared to previous years [8,9]. Influenza and respiratory syncytial virus (RSV) sharply declined during the winter of 2020/2021 and this was largely attributed to a decline in transmission of these viruses [10–12]. Conversely, certain conditions, like those related to mental health, were predicted to increase in incidence during pandemic shutdowns. Initial studies of self-reported depression and anxiety recorded during April 2020 supported this hypothesis, yet there has been mixed evidence on how this has translated to mental health-related hospitalizations [13–15]. Analyses of trends in non-COVID-19 disease during the pandemic period have largely focused on specific subsets of health conditions and have not taken a comprehensive view of disease-specific trends in relation to overall trends in hospitalization through multiple waves of SARS-CoV-2. A systematic analysis of diagnoses prior to and during the pandemic period is necessary to provide the comprehensive estimates of disease burden and to aid in hospital preparation and resource allocation during subsequent waves of COVID-19 [16,17].

In this study, we leverage a large national healthcare billing database to estimate relative changes in the incidence of hospitalizations by diagnostic category as the COVID-19 pandemic emerged and progressed through multiple waves of SARS-COV-2 variants. Our examination identifies broad patterns, which inform hypotheses regarding the influence of NPIs, diagnostic testing, and healthcare seeking on the reported incidence of disease. The results of this research provide a unique high-level understanding of the role of disruptions to healthcare and social interactions on multiple illnesses and can help to interpret reported trends in surveillance and hospitalization data.

## Methods

### COVID-19 Research Database

Data were provided by the *COVID-19 Research Database* (C19RDB), which is a pro-bono public-private consortium that collects and aggregates de-identified claims data, such as healthcare claims data. For this study, we used a health insurance claims (i.e. billing) database that included identifiers for healthcare setting (inpatient, outpatient), primary diagnostic code, secondary diagnostic codes, billing site location (state), date of service, Current Procedural Terminology codes, age and gender, among other billing details. Reporting to this database was continually updated until September 2021, leading to variation in total claims per month as additional payers were incorporated over time. This database specifically captures billing data at the level of individual hospitalizations, along with associated demographic, diagnostic, and procedure code information. Information on the total population within each insurance payer (i.e. payees) was not available, limiting our ability to accurately assess a population denominator from which hospitalizations arose. This research has been approved by the Georgetown University Institutional Review Board.

### Data Cleaning

We limited the data to inpatient records due to large and unexplained fluctuations in the volume of outpatient data entering the database in early 2019. Only hospitalizations with a corresponding procedure code were included for analysis. Based on anonymized patient identification numbers, entries were deduplicated to retain only the first hospitalization record within a 30-day rolling period for each patient. The month and year of diagnosis used for analysis was the “service from date” when available, otherwise “service to date” or “admission date” or “billing statement from” date was used (in order of first availability). “Service from date” was present in the majority (>90%) of records and the primary source for date information. Hospitalizations for COVID-19 were excluded from all analyses if a code for COVID-19 (ICD-10 “U07.1 – COVID-19” or “J12.82 – pneumonia due to SARS-associated coronavirus” or “M35.81 – COVID-19 related multisystem inflammatory syndrome” or “B97.2* - coronavirus as the cause of disease elsewhere classified”) was present, in order to properly assess trends in non-COVID-19 illnesses [18]. The final study period was January 1, 2017 to June 31, 2021 (**Supplementary Figure S1**).

The outcome of interest was the primary (i.e. first) diagnostic field recorded for each visit as categorized by the International Classification of Diseases-Clinical Modification (ICD-10-CM) codes. We included only the primary diagnostic code for each hospitalization in order to limit the influence of diagnoses not directly related to admission [19]. Primary diagnostic codes for all patients were grouped by ICD-10 chapter (2016 edition of ICD-10-CM), subchapter, month, and year. These chapters are organized according to illness type and/or body system affected, totaling 22 chapters, of which diagnostic codes falling into chapters 1 through 19 were assessed (excluding chapters for external morbidity, special purposed codes, and health services contact codes) (**Supplementary Table S1**). Subchapter groupings were created according to categories defined by the World Health Organization (WHO) and were primarily organized by type of infectious agent identified, type of illness, or part of body affected in addition to codes for “unspecified” or “unknown” illness [20]. The 19 chapters could be subdivided into 236 subchapters, but subchapters with fewer than 5000 diagnoses total between 2017 and 2019 were excluded from all analyses (47 exclusions) (**Supplementary Table S2**).

### Analysis

The aim of this analysis was to provide a monthly estimate of the incidence rate ratio (IRR), defined as the observed number of monthly hospitalizations divided by the expected monthly hospitalizations, for each diagnostic chapter and subchapter of interest. Estimates were generated using Poisson regression models fit to the monthly count of diagnostic codes within each ICD-10 chapter (or subchapter) from the pre-pandemic period (January 1, 2017 – December 31, 2019) to estimate the number of hospitalizations per month had the pandemic not occurred. Time series for each chapter and subchapter were analyzed independently. All models adjusted for time trends (linear trend) and yearly seasonality (using sine and cosine harmonic terms). While we did not have a true population denominator to use as an offset term in the mode, due to a lack of information on payer population or hospital catchment areas, we estimated the size of the active patient population as the number of unique individuals with a recorded hospitalization per year for any cause. For the hospitalizations occurring between 2017 and 2019, the offset was the total number of unique individuals present in the same calendar year. Because there was a generally increasing trend in the number of individuals included in this database, the offset for 2020 and 2021 was estimated by fitting linear and seasonal trends to the monthly counts of unique individuals in the study prior to March 2020 and extrapolating these trends to the period from March 2020-June 2021. Prediction intervals for the final IRR estimates were calculated for each time point through a two-stage simulation using Monte Carlo resampling that accounted for parameter uncertainty and observation uncertainty [21,22].

Due to the numerous subchapters included in this analysis (N = 180), we implemented a hierarchical clustering algorithm (applying Ward’s minimum variance method) to systematically group diagnoses that shared temporal trends in IRRs during the period between Jan 2020 and Jun 2021. The optimal number of clusters (k) was chosen via the “elbow method”. This method attempts to maximize the variance (within-cluster sum of squares) explained by the clustering and minimize the number of clusters thereby excluding an increasing number of clusters that only provide a marginal gain in explained variance. Data queries and analyses were run in SQL and R version 4.0.2 using the packages data.table, DBI, dplyr, ggplot2, glue, gridExtra, lubridate, and readr.

## Results

The total number of unique hospitalized individuals ranged from 2.8 million to 3.1 million per year between 2017 and 2020. Insurance payers covered patients from all fifty states and Puerto Rico. Between January 2017 and December 2019, the monthly number of unique hospitalizations (excluding repeat admissions occurring within a 30-day period and COVID-19 admissions) ranged from ∼300,000 to ∼400,000. In 2020, this number dropped from 455,000 unique hospitalizations in January to 263,000 in March and then declined again to 151,000 encounters in April 2020, before rebounding moderately to 213,000 in June.

### Trends in broad disease classes

All ICD10 chapters experienced relative increases in incidence of hospitalizations during the pre-pandemic months of January and February 2020, with IRRs ranging from 0.96 to 1.21 in Jan 2020 (**Figure 1; Supplemental Table S3**), likely reflecting database reporting changes at the start of the new calendar year. Beginning in March 2020, the incidence of hospitalization among all diagnostic chapters exhibited a moderate decrease. For most of chapters, the greatest declines in incidence occurred during April 2020 (**Supplementary Figure S2)**. This was especially true for *diseases of the eye* and *diseases of the ear* while the smallest changes in hospitalizations were found among conditions classified within *mental health, perinatal period*, and *pregnancy, childbirth, and puerperium* chapters. The incidence of hospitalizations for *respiratory disease* was lower than expected for all months following April 2020, reaching the greatest relative decline in January 2021, when seasonal hospitalizations would typically be increasing (IRR 0.29; 95% CI: 0.28, 0.29).

**Figure 1.**
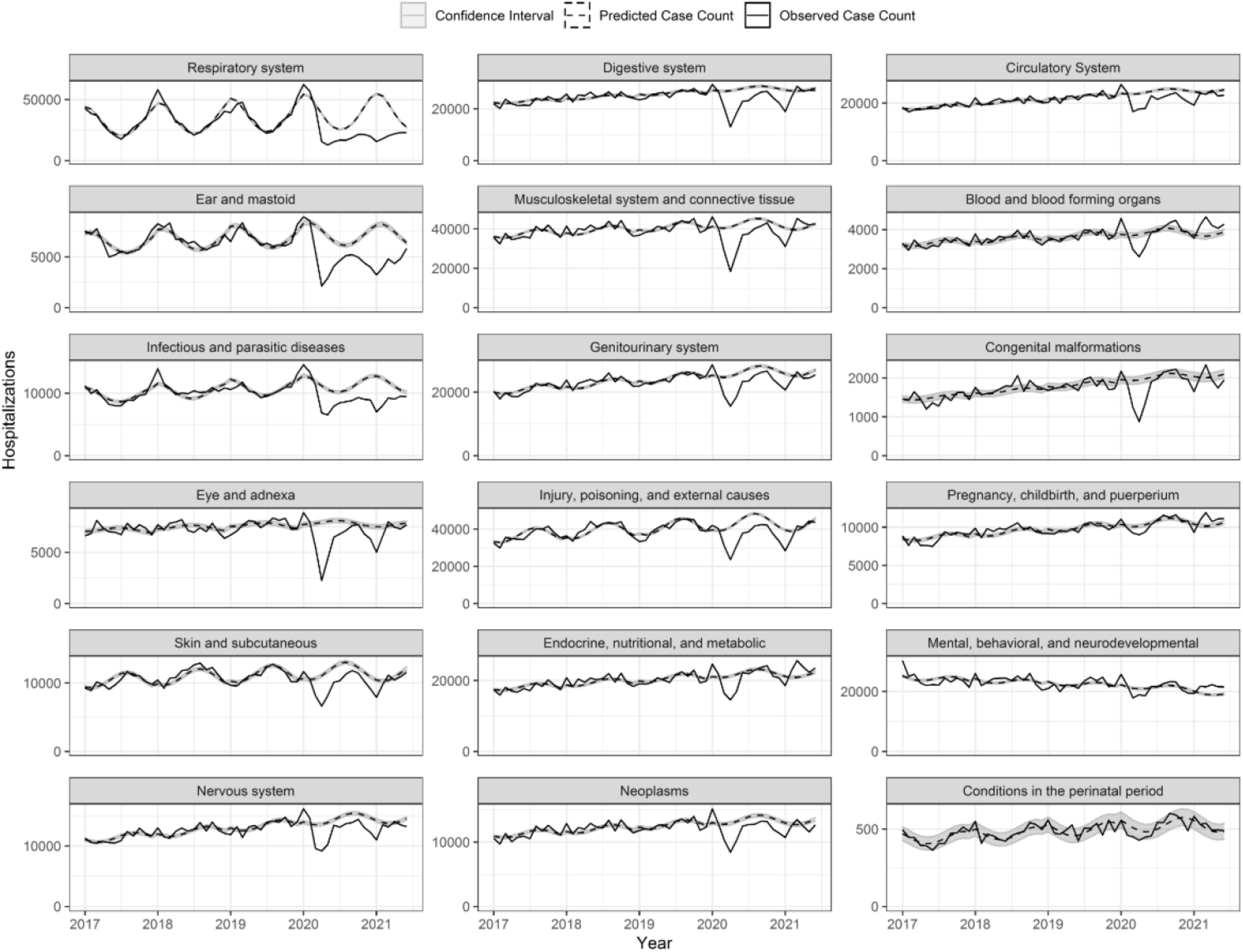
Time series of observed hospitalizations per diagnostic chapter by month and year (solid line) with model predicted case counts (dashed line) and confidence intervals (grey).

### Clustering of trends within diagnostic subchapters

A cluster analysis revealed 4 distinct groupings of ICD-10 subchapters (**Supplemental Figures S3-S4)**. Subchapters grouped in cluster A shared a sharp relative decrease in hospitalizations in April 2020 that continued to have below-expected rates throughout the remainder of the 2020. The average IRR of this cluster was 0.46 (range: 0.11, 1.09) between March 2020 and June 2021. These subchapters included diagnoses for Pneumonia & Influenza (P&I) (J09 -J18), Middle Ear (H65-H75), Intestinal Infectious diseases (A00-A09), Conjunctivitis (H10-H11), Viral skin and mucous infections (B00-B09), Biomechanical lesions (M99), Acute lower respiratory infection (LRI), and Acute upper respiratory infection (URI) (**Figure 2A**). The incidence of P&I in January and February 2021 was 89% (95%CI: 88.7%, 90.4%) lower than what would be expected given predicted trends in incidence from previous years and seasonal trends (**Supplemental Table S4**). There were just 9,427 unique diagnoses for P&I between Oct 2020 and Mar 2021 compared to 61,085 during the same months of 2019. The timing and intensity of the decline in incidence varied by subchapter within this cluster, with most subchapters experiencing the greatest decline in April 2020 or January 2021. Codes for *biomechanical lesions* (not elsewhere classified) were unique in that their lowest IRR was recorded in Nov 2020 of 0.20 (0.17, 0.23). While diagnoses for biomechanical lesions decreased during the summer of 2020 compared to what was expected, the magnitude of the declines for biomechanical lesions showed much greater uncertainty compared to the estimates for infectious diseases, reflecting the small case counts for primary diagnoses of this subchapter within the dataset. The only diagnostic subchapters from this cluster A to approach pre-pandemic incidence levels (i.e. RR approaching 1.0) were *intestinal infectious diseases, acute URI*, and *acute LRI* in May and June of 2021.

**Figure 2A.**
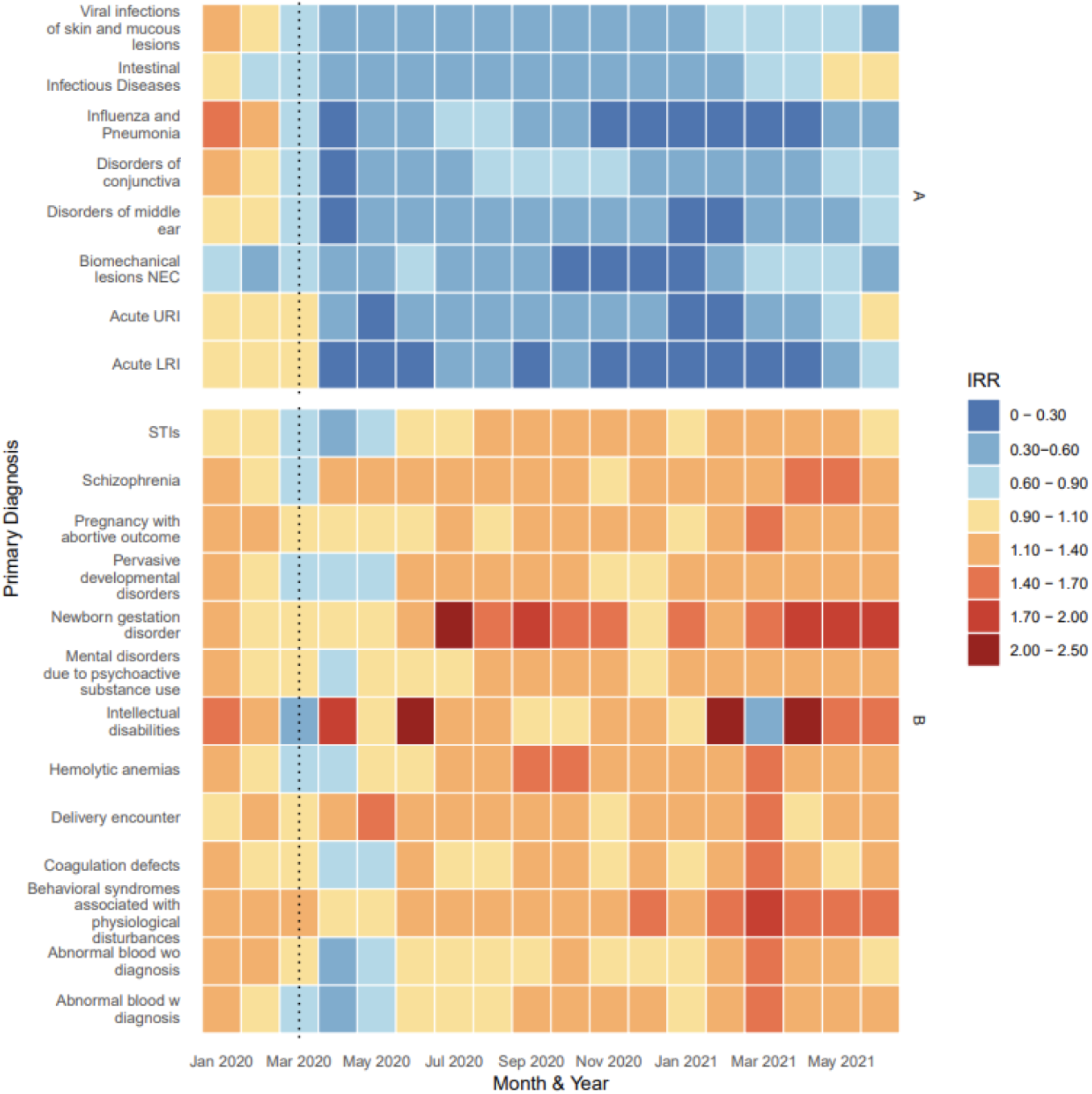
Estimated IRR per subchapter and month grouped according to hierarchical clustering, Jan 2020 to Jun 2021

In contrast to cluster A, cluster B had little to no decrease in hospitalizations during March, April or May of 2020. The average IRR in this cluster remained below expected levels only for the months of March, April, and May 2020, prior to returning to and exceeding pre-pandemic levels for the remainder of the study period. Diagnoses within this cluster included: *Sexually Transmitted Infections* (*STI*s) (A50-A64), *intellectual disabilities* (F70-F79), *psychological disturbances* (F50-F59), and *coagulation defects* (D65-D69) in addition to ICD-10 codes related to pregnancy such as *delivery encounter* (O80-O82), *pregnancy with an abortive outcome* (O00-O08), and *newborn gestation disorder* (P05-P08). The average IRR of this cluster between March 2020 and June 2021 was 1.20 (range: 0.46, 2.20).

Cluster C included diagnoses that decreased in incidence during March, April, and May of 2020, and well as January of 2021, but hovered around pre-pandemic estimates of incidence for the remainder of the study period (**Figure 2B**). This grouping included diagnoses for *hypertension* (I10-I16), *malnutrition* (E40-E46), *diseases of appendix* (K35-K38), *diabetes* (E08-E13), and *neoplasms* of multiple systems (e.g. respiratory, CNS, reproductive organs, soft tissue, breast, digestive organs, etc.). The average IRR of this cluster between March 2020 and June 2021 was 0.95 (range: 0.12, 1.85). Subchapters of cluster C demonstrated an average IRR < 1.0 during 2020 but incidence began to increase gradually during 2021. Diseases of the lens, which experienced the largest drop in incidence during April 2020 (IRR: 0.12; 95% PI: 0.12, 0.13) was included in this cluster as it quickly returned to its pre-pandemic incidence range by June 2020.

**Figure 2B.**
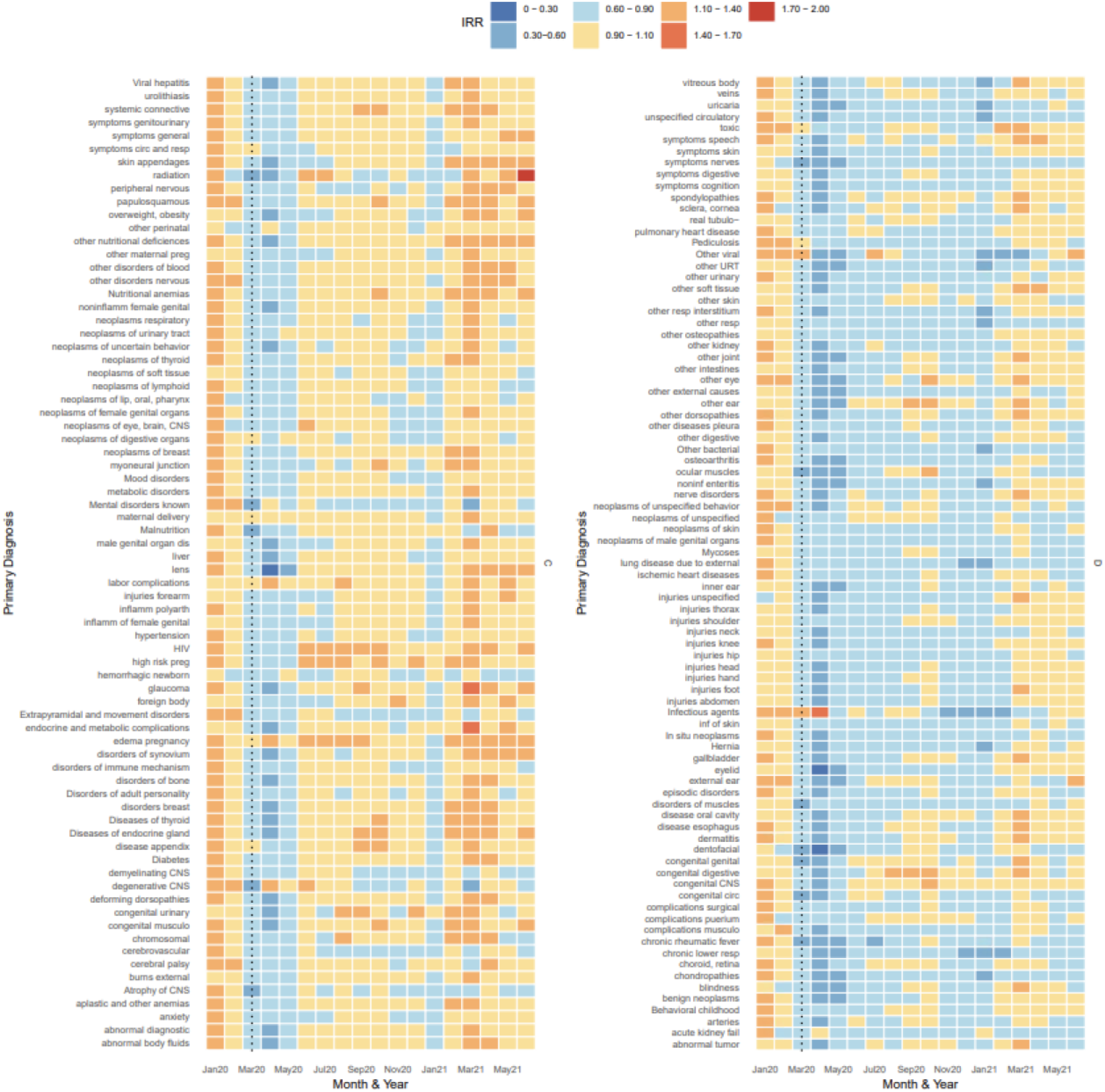
Estimated IRR per subchapter and month grouped according to hierarchical clustering (clusters C and D), Jan 2020 to Jun 2021

Cluster D exhibited a relative decrease in hospitalizations during March, April, and May 2020 and continued to exhibit lower-than-expected incidence during the remainder of 2020 and early 2021 with occasional months near baseline, particularly during Aug, Sept, Oct (**Figure 2B)**. This included diagnoses such as *pulmonary heart disease* (I26-I28), *ischemic heart diseases* (I20-I25), *injuries* of multiple areas of the body (thorax, shoulder, neck, knee, hip, head, hand, foot)(S00-S99), *chronic lower respiratory disease* (J40-J47), *chronic rheumatic fever* (I05-I09), and *neoplasms of the skin* (C43-C44). On average, the diseases in these clusters had an IRR of 0.83 (range: 0.29, 1.45) between March and June 2021.

## Discussion

The COVID-19 pandemic has profoundly disrupted healthcare and disease dynamics in the United States. The wealth of data provided by this healthcare insurance billing database allowed for a novel, large scale examination of these disruptions for different disease categories. The vast majority of diagnostic categories experienced a decline in relative incidence of hospitalizations during April and May of 2020, while the timing at which incidence returned to pre-pandemic levels varied by diagnostic chapter, and some chapters have yet to reach pre-pandemic levels of incidence. The unique and robust data leveraged in this analysis allowed for exploration of trends in hospitalizations that we hypothesize might be due to a range of pandemic changes in healthcare utilization, lifestyle changes, and sanitation practices.

Our analysis was able to confirm findings from prior, smaller studies of illness using a national database, and was thus able to provide more substantial estimates of relative incidence. Hospitalizations with a primary diagnostic code for infectious respiratory disease (e.g. influenza, acute lower respiratory infections) and otitis media (the main contributor to the “middle ear diagnosis” subchapter) clustered in our analysis given their shared dramatic decrease in IRR during April and May 2020 and their uncharacteristically low relative incidence during fall/winter 2020/2021. These results substantiate prior studies of influenza, RSV and other upper and lower respiratory infections during the winter 2020/2021 season [10,23]. NPIs are believed to have reduced influenza transmission during the 2020/2021 influenza season while also contributing to the decline in mumps and other respiratory infections in some countries [12,23–25]. While the decline in influenza could be partially attributed to a decrease in the number of specimens sent for respiratory panel testing, as evidenced by an 61% decrease in US specimens sent for testing in 2020, studies have shown that the positivity rate among specimens sent had also declined, approximately 98%, during winter 2020/2021 [11]. Our finding that middle ear infections (otitis media, OM) remained low following pandemic lockdowns is supported by multiple studies in Milan where authors identified reductions in OM among OM-prone children of ∼80% between Feb and May 2020 compared to the same time period in 2019 [26,27].

In contrast to the subchapters with large declines in incidence, the cluster of subchapters with increased rates (cluster B), which included diagnoses for *behavioral syndromes associated with physiological disturbances* (F50-F59), *hemolytic anemias* (D55-D59), and *intellectual disabilities* (F70-F79). Interestingly, these illnesses did not appear to have a clear shared mechanism to explain the pandemic-period trends. Potential drivers of the sporadic *increases* in disease incidences during the pandemic period include: 1) changes in the underlying population covered by the insurance payers included in our analysis or changes in which hospitals (or types of hospitals that) are within the insurance service areas, 2) diagnoses that are a direct result of a prior SARS-CoV-2 infection, and 3) mechanisms that cannot be elucidated using this dataset such as diagnostic screening, test ordering, and the prioritization of hospitalization vs outpatient care settings. Prior research has revealed that patients with intellectual disabilities are more likely to contract COVID-19 and more likely to require hospitalization compared to those without an established intellectual disability [28]. Our analysis excluded any diagnoses for COVID-19, but it is possible that the elevated incidence of hospitalizations where the primary diagnosis is for intellectual disability could be linked to the downstream effects of a prior COVID-19 infection or other pandemic-related changes in care for members of this population.

There has been limited research assessing the impact of the pandemic on behavioral syndromes associated with physiological disturbances (e.g. eating and sleep disorders); however, researchers at a children’s hospital did describe an increase in restrictive eating disorders during the first year of the pandemic [29]. Contrary to our results, psychiatric hospitalizations in Brazil decreased between March and August 2020 by 33%, demonstrating the complex relationship between physiological disturbances, schizophrenia, and behavioral syndromes incidence and hospitalization incidence [30]. Interestingly, schizophrenia is noted as a risk factor for COVID-19 mortality, but our study was limited by its inability to explore mortality information [31]. An increase in hemolytic anemias during the pandemic period was also identified; however, further research is needed to validate our initial findings given that no apparent mechanism for this trend could be elucidated from this data [32,33]. The development of autoimmune hemolytic anemias in conjunction with SARS-COV-2 infection have been reported; however, our analysis excluded any such hospitalizations so the mechanism responsible for this increase in our data is unknown [34,35].

The ICD-10 chapter for *Pregnancy, Childbirth, and Puerperium* (O00-O9A) only experienced an estimated decrease in incidence of 11% (95% PI: 9%,13%) in April of 2020. ICD-10 codes for labor and delivery would not be expected to drop in the early months of the pandemic as the number of expected births could not be influenced by pandemic-related changes in family planning. However, birth outcomes could be influenced by the level of prenatal care and pregnancy-related stress as the pandemic continued into late 2020 and early 2021. Throughout the study period, incidence of *newborn gestation disorder* was aberrant, vacillating between an estimate IRR of 0.68 and 2.20, and did not appear to clearly alter in relation to COVID-19 pandemic changes in health care. The increase in incidence during 2020 and 2021 for these diagnostic codes could be a product of an otherwise increasing trend in data reporting to the database or a true reflection of adverse newborn outcomes related to the COVID-19 pandemic.

Cluster D included diagnoses that lacked specificity, such as “*symptoms for*….”, “*other disorders of*….”, and “*complications of*…” diagnoses, and may not be commonly coded within the primary diagnosis field. Hospitalizations for injuries clustered in this category as well, with most experiencing a decline in incidence until Jan 2021. These findings are supported by a few studies detailing trauma center admissions, with large declines among injuries due to road traffic conditions which constitute a large portion of trauma admissions [36,37]. Conversely, increases in certain types of severe injuries (i.e. self-inflicted injuries) have been reported in some emergency settings during the early period of the pandemic, including “penetrating injuries” [38,39]. As our analysis relies on inpatient data, we are likely only seeing a small portion (and severe enough to require an inpatient hospitalization) of the total injuries that were presenting during this time.

In contrast to diagnoses that had a slow rebound in incidence of hospitalizations during 2020, like those in cluster D, hospitalizations for neoplasms (cluster C) had a short-lived decline between March and June 2020, followed by an extended period of time hovering around pre-pandemic levels. The largest decrease among all neoplasm diagnoses was among benign neoplasms in April 2020 (−77%, PI: -76%, -78%). Neoplasms the skin, thyroid, and male genital track also had a decrease in hospitalizations of 30% or more in April 2020. The most likely explanation for the fluctuations in neoplasm diagnoses are disruptions to cancer screenings and subsequent diagnoses resulting from healthcare access limitations during the early pandemic period followed by an influx of patients in need of delayed cancer screenings and referrals [40–43]. Cluster C, as a whole, could be characterized as declining in relative incidence in correspondence with waves of COVID-19 occurring in the US, primarily March and April of 2020 and January 2021. This supports the hypothesis that these hospitalizations are primarily those that could be delayed during peak COVID-19 transmission cycles.

### Strengths and Limitations

Due to the passive nature of data reporting to the healthcare encounter billing clearinghouse and the differential reporting by the insurance payer, different payers may contribute to relative increases or decreases in disease rates in a manner that might not be reflective of true trends present across the US during the study period. Alternative explanations for the relative increases in disease diagnoses include payer catchment area redefinitions (and sudden inclusions of specialty facilities), reimbursement policies, and primary vs secondary diagnostic field coding practices. Our analysis was limited to primary diagnostic codes, thus the full range of diagnoses made during the study period were not captured and it is possible that IRR trends may vary if the full breadth of secondary codes were included for analysis. This study was also limited to inpatient data; therefore, we are unable to discern if relative increases or decreases in hospitalization may be linked to a shift in treatment location (i.e. cases being treated in an outpatient setting that would otherwise be hospitalized or vice versa). This phenomenon has been noted among cases of appendicitis such that more cases were treated with non-operative management prior to delayed operative treatment during the early pandemic period [44,45].

For most diagnostic chapters and subchapters, there was an increase in hospitalizations in Jan and Feb 2020 which indicated a fluctuation in the billing data reported to the database used for this analysis. This complicates the interpretation of elevated incidence rates among certain diagnostic subchapters during the pandemic period as it may be a reflection of the overall influx of data to this clearinghouse that began at the start of the 2020 calendar year. Additionally, some diagnostic chapters had a low frequency of events, particularly during the pandemic period, such that we could not obtain precise estimates of their monthly IRR.

Our analysis is strengthened by the large study population size (28 million hospitalizations) and the ability to investigate rare disorders that may otherwise not be captured in an analysis of disease trends. Our ability to access multiple years of data prior to the pandemic also strengthens our confidence in estimates of relative incidence during the pandemic period. Examination of billing data allowed this study to overcome limitations in disease reporting due to COVID-19 related disruptions, as has been demonstrated for the Nationally Notifiable Disease Surveillance System (NNDSS) data [8].

## Conclusion

There were substantial changes in hospitalization patterns across many different disease categories during the early part of the COVID-19 pandemic, some of which persisted for at least a year. The broad trends identified here point to a number of hypotheses about the mechanisms driving these trends. Declines in infectious respiratory disease may likely be a result of NPI use, social distancing, and changes to daily life that altered contact patterns. Meanwhile, this research has highlighted potential areas for further investigation by experts in the field, such as trends in hospitalizations for intellectual disabilities, newborn gestation disorders, and mental health disorders. Understanding these patterns will be crucial to minimize disruptions to healthcare during future major health events.

## Data Availability

The data that support the findings of this study are available from COVID-19 Research Database consortium, but restrictions apply to the availability of these data, which were used under license for the current study, and so are not publicly available. Data are however available from the authors upon reasonable request and with permission of COVID-19 Research Database consortium.

## Author Contributions

Concept and design: SB, DMW

Acquisition, analysis, and data interpretation: SB, KC, DMW, CZ

Statistical analysis: KC, DMW

Drafting of the manuscript: SB, KC, DMW

Manuscript revision and final approval: SB, KC, DMW, CZ

## Acknowledgements

*The data, technology, and services used in the generation of these research findings were generously supplied pro bono by the COVID-19 Research Database partners, who are acknowledged at https://covid19researchdatabase.org/. We would also like to thank Dominic Copper-Wooten for his contributions regarding SQL querying*.

## Data Availability

The data that support the findings of this study are available from *COVID-19 Research Database consortium*, but restrictions apply to the availability of these data, which were used under license for the current study, and so are not publicly available. Data are however available from the authors upon reasonable request and with permission of *COVID-19 Research Database consortium*.

## Figures and Tables

**Supplemental Figure S1.**
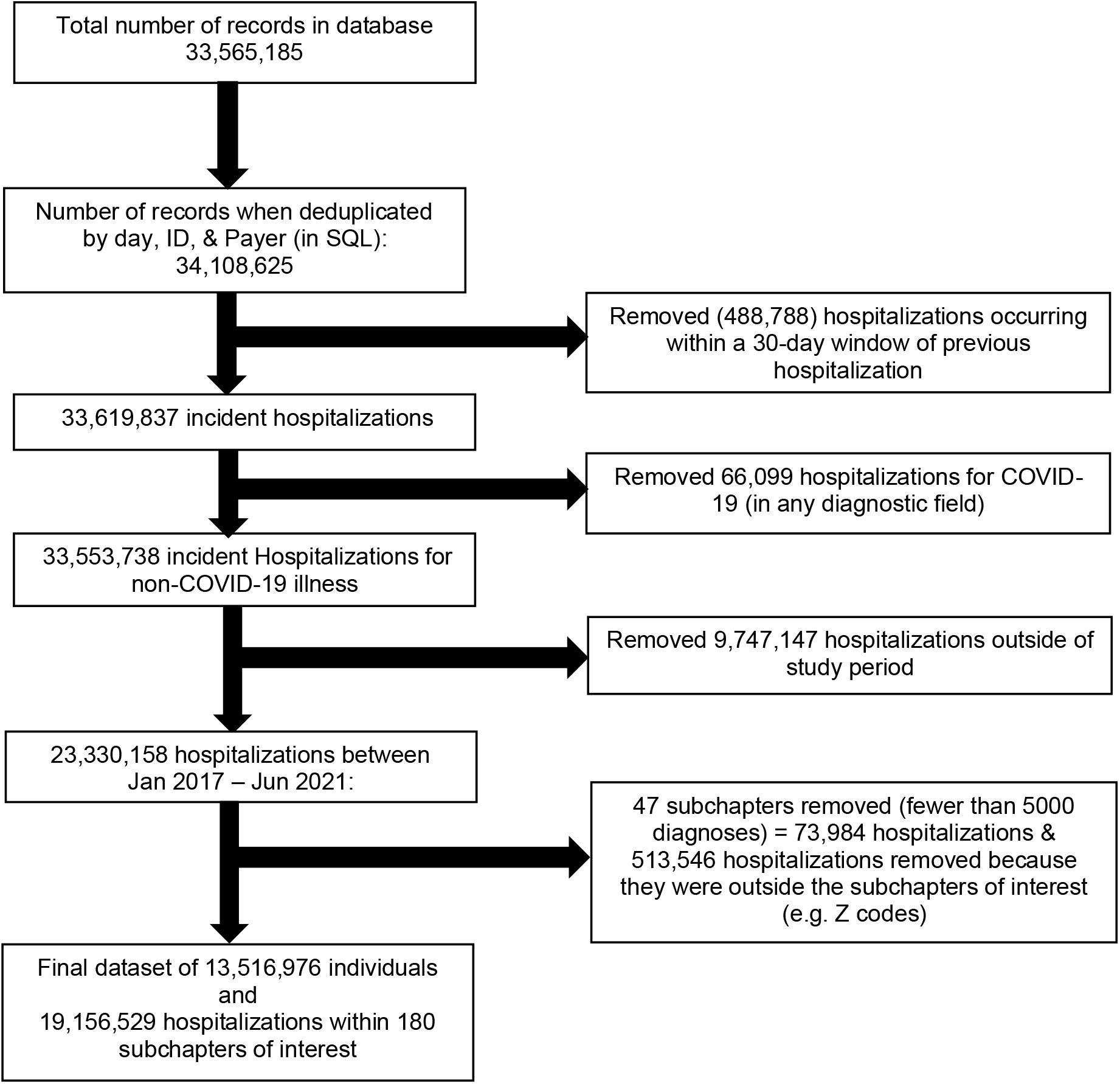
Flowchart illustrating the data subsetting process for the COVID-19 Research Database inpatient data, Jan 1, 2017 – Jun 31, 2021

**Supplementary Table S1.** Categorization of ICD-10 Chapters and diagnostic codes

| Chapter No. | ICD-10 Code Range | Description |
| --- | --- | --- |
| 1 | A00 – B99 | Certain infectious and parasitic diseases |
| 2 | C00 – D49 | Neoplasms |
| 3 | D50 – D89 | Diseases of the blood and blood-forming organs and certain disorders involving the immune mechanism |
| 4 | E00 – E89 | Endocrine, nutritional and metabolic diseases |
| 5 | F01 – F99 | Mental, behavioral and neurodevelopmental disorders |
| 6 | G00 – G99 | Diseases of the nervous system |
| 7 | H00 – H59 | Diseases of the eye and adnexa |
| 8 | H60 – H95 | Diseases of the ear and mastoid process |
| 9 | I00 – I99 | Diseases of the circulatory system |
| 10 | J00 – J99 | Diseases of the respiratory system |
| 11 | K00 – K95 | Diseases of the digestive system |
| 12 | L00 – L99 | Diseases of the skin and subcutaneous tissue |
| 13 | M00 – M99 | Diseases of the musculoskeletal system and connective tissue |
| 14 | N00 – N99 | Diseases of genitourinary system |
| 15 | O00 – O9A | Pregnancy, childbirth and the puerperium |
| 16 | P00 – P96 | Certain conditions originating in the perinatal period |
| 17 | Q00 – Q99 | Congenital malformations, deformations and chromosomal abnormalities |
| 18 | R00 – R99 | Symptoms, signs, and abnormal clinical laboratory findings, not elsewhere classified |
| 19 | S00 – T88 | Injury, poisoning and certain other consequences of external causes |

**Supplementary Table S2.** Excluded subchapters (fewer than 500 diagnoses between Jan1 2017-June 2021)

|  |  |
| --- | --- |
| Acute rheumatic fever (I00-I02) | Infectious arthropathies (M00-M02) |
| Arthropod-borne (A90-A99) | Infectious and parasitic sequelae (B90-B94) |
| Autoinflammatory syndromes (M04) | Inflammatory CNS (G00-G09) |
| Birth trauma (P10-P15) | Injuries multiple (T07) |
| Bullous disorders (L10-L14) | Necrotic lower respiratory (J85-J86) |
| Burns internal (T26-T28) | Neoplasms of bone (C40-C41) |
| Burns multiple (T30-T32) | Newborn complications (P00-P04) |
| Certain zoonotic bacterial (A20-A28) | Optic nerve (H46-H47) |
| Cleft lip (Q35-Q37) | Other chlamydiae (A70-A74) |
| Complications of endocrine system (E36) | Other heart disease (I30-I5A) |
| Complications genitourinary (N99) | Other human herpesviruses (B10) |
| Complications puerperium (O85-O92) | Other infectious (B99) |
| Complications trauma (T79) | Other musculoskeletal (M95) |
| Complications skin (L76) | Other newborn (P84) |
| Complications spleen (D78) | Other spirochetal (A65-A69) |
| Complications respiratory (J95) | Periprosthetic fracture (M97) |
| Congenital malformations respiratory (Q30-Q34) | Diseases of peritoneum (K65-K68) |
| Digestive newborn (P76-P78) | Protozoal disease (B50-B64) |
| Disorders of glucose and pancreas (E15-E16) | Respiratory-cardio perinatal (P19-P29) |
| Ear complications (H95) | Rickettsioses (A75-A79) |
| Endocrine newborn (P70-P74) | Temperature newborn (P80-P83) |
| Eye complications (H59) | Tuberculosis (A15-A19) |
| Frostbite (T33-T34) | Unknown mortality (R99) |
| Glomerular diseases (N00-N08) | Unspecified mental (F99) |
| Helminthiasis (B65-B83) | Viral prion, infections of CNS (A80-A89) |
|  | Ill-defined and unknown mortality (R99) |

**Supplementary Table S3.**
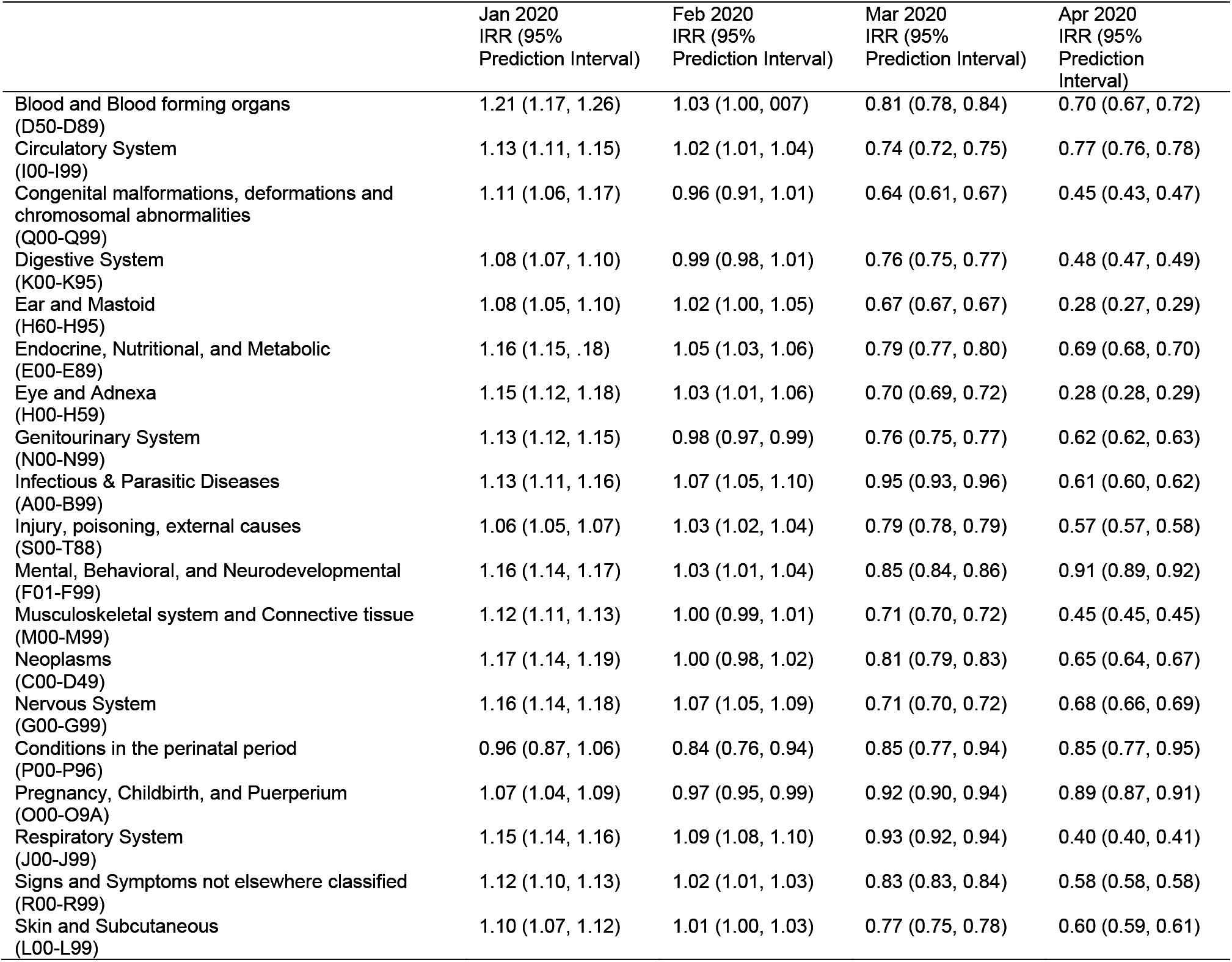
Estimated IRR and 95% Prediction Interval of select ICD10 chapters between Jan and Apr 2020

|  | Jan 2020<br>IRR (95%<br>Prediction Interval) | Feb 2020<br>IRR (95%<br>Prediction Interval) | Mar 2020<br>IRR (95%<br>Prediction Interval) | Apr 2020<br>IRR (95%<br>Prediction<br>Interval) |
| --- | --- | --- | --- | --- |
| Blood and Blood forming organs<br>(D50-D89) | 1.21 (1.17, 1.26) | 1.03 (1.00, 1.07) | 0.81 (0.78, 0.84) | 0.70 (0.67, 0.72) |
| Circulatory System<br>(I00-I99) | 1.13 (1.11, 1.15) | 1.02 (1.01, 1.04) | 0.74 (0.72, 0.75) | 0.77 (0.76, 0.78) |
| Congenital malformations, deformations and<br>chromosomal abnormalities<br>(Q00-Q99) | 1.11 (1.06, 1.17) | 0.96 (0.91, 1.01) | 0.64 (0.61, 0.67) | 0.45 (0.43, 0.47) |
| Digestive System<br>(K00-K95) | 1.08 (1.07, 1.10) | 0.99 (0.98, 1.01) | 0.76 (0.75, 0.77) | 0.48 (0.47, 0.49) |
| Ear and Mastoid<br>(H60-H95) | 1.08 (1.05, 1.10) | 1.02 (1.00, 1.05) | 0.67 (0.67, 0.67) | 0.28 (0.27, 0.29) |
| Endocrine, Nutritional, and Metabolic<br>(E00-E89) | 1.16 (1.15, 1.18) | 1.05 (1.03, 1.06) | 0.79 (0.77, 0.80) | 0.69 (0.68, 0.70) |
| Eye and Adnexa<br>(H00-H59) | 1.15 (1.12, 1.18) | 1.03 (1.01, 1.06) | 0.70 (0.69, 0.72) | 0.28 (0.28, 0.29) |
| Genitourinary System<br>(N00-N99) | 1.13 (1.12, 1.15) | 0.98 (0.97, 0.99) | 0.76 (0.75, 0.77) | 0.62 (0.62, 0.63) |
| Infectious & Parasitic Diseases<br>(A00-B99) | 1.13 (1.11, 1.16) | 1.07 (1.05, 1.10) | 0.95 (0.93, 0.96) | 0.61 (0.60, 0.62) |
| Injury, poisoning, external causes<br>(S00-T88) | 1.06 (1.05, 1.07) | 1.03 (1.02, 1.04) | 0.79 (0.78, 0.79) | 0.57 (0.57, 0.58) |
| Mental, Behavioral, and Neurodevelopmental<br>(F01-F99) | 1.16 (1.14, 1.17) | 1.03 (1.01, 1.04) | 0.85 (0.84, 0.86) | 0.91 (0.89, 0.92) |
| Musculoskeletal system and Connective tissue<br>(M00-M99) | 1.12 (1.11, 1.13) | 1.00 (0.99, 1.01) | 0.71 (0.70, 0.72) | 0.45 (0.45, 0.45) |
| Neoplasms<br>(C00-D49) | 1.17 (1.14, 1.19) | 1.00 (0.98, 1.02) | 0.81 (0.79, 0.83) | 0.65 (0.64, 0.67) |
| Nervous System<br>(G00-G99) | 1.16 (1.14, 1.18) | 1.07 (1.05, 1.09) | 0.71 (0.70, 0.72) | 0.68 (0.66, 0.69) |
| Conditions in the perinatal period<br>(P00-P96) | 0.96 (0.87, 1.06) | 0.84 (0.76, 0.94) | 0.85 (0.77, 0.94) | 0.85 (0.77, 0.95) |
| Pregnancy, Childbirth, and Puerperium<br>(O00-O9A) | 1.07 (1.04, 1.09) | 0.97 (0.95, 0.99) | 0.92 (0.90, 0.94) | 0.89 (0.87, 0.91) |
| Respiratory System<br>(J00-J99) | 1.15 (1.14, 1.16) | 1.09 (1.08, 1.10) | 0.93 (0.92, 0.94) | 0.40 (0.40, 0.41) |
| Signs and Symptoms not elsewhere classified<br>(R00-R99) | 1.12 (1.10, 1.13) | 1.02 (1.01, 1.03) | 0.83 (0.83, 0.84) | 0.58 (0.58, 0.58) |
| Skin and Subcutaneous<br>(L00-L99) | 1.10 (1.07, 1.12) | 1.01 (1.00, 1.03) | 0.77 (0.75, 0.78) | 0.60 (0.59, 0.61) |

**Supplementary Figure S2.**
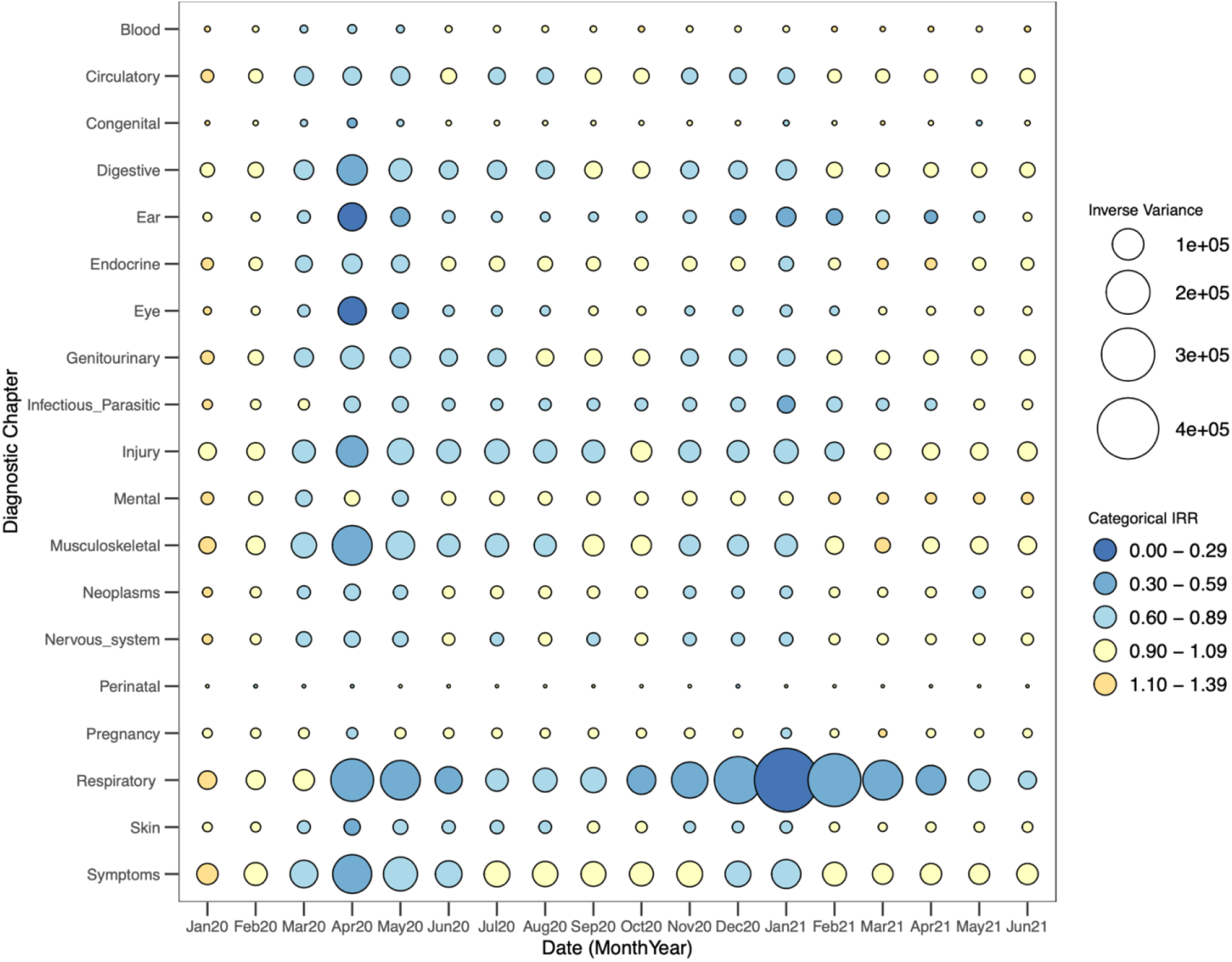
Incidence Rate Ratio (IRR) per Diagnostic Chapter for the months between January 2020 and June 2021S Each bubble is colored according to IRR and the diameter of the bubble is proportional to the inverse of the IRR variance estimate (i.e. a larger bubble signifies greater confidence in the estimate).

**Supplemental Figure S3:**
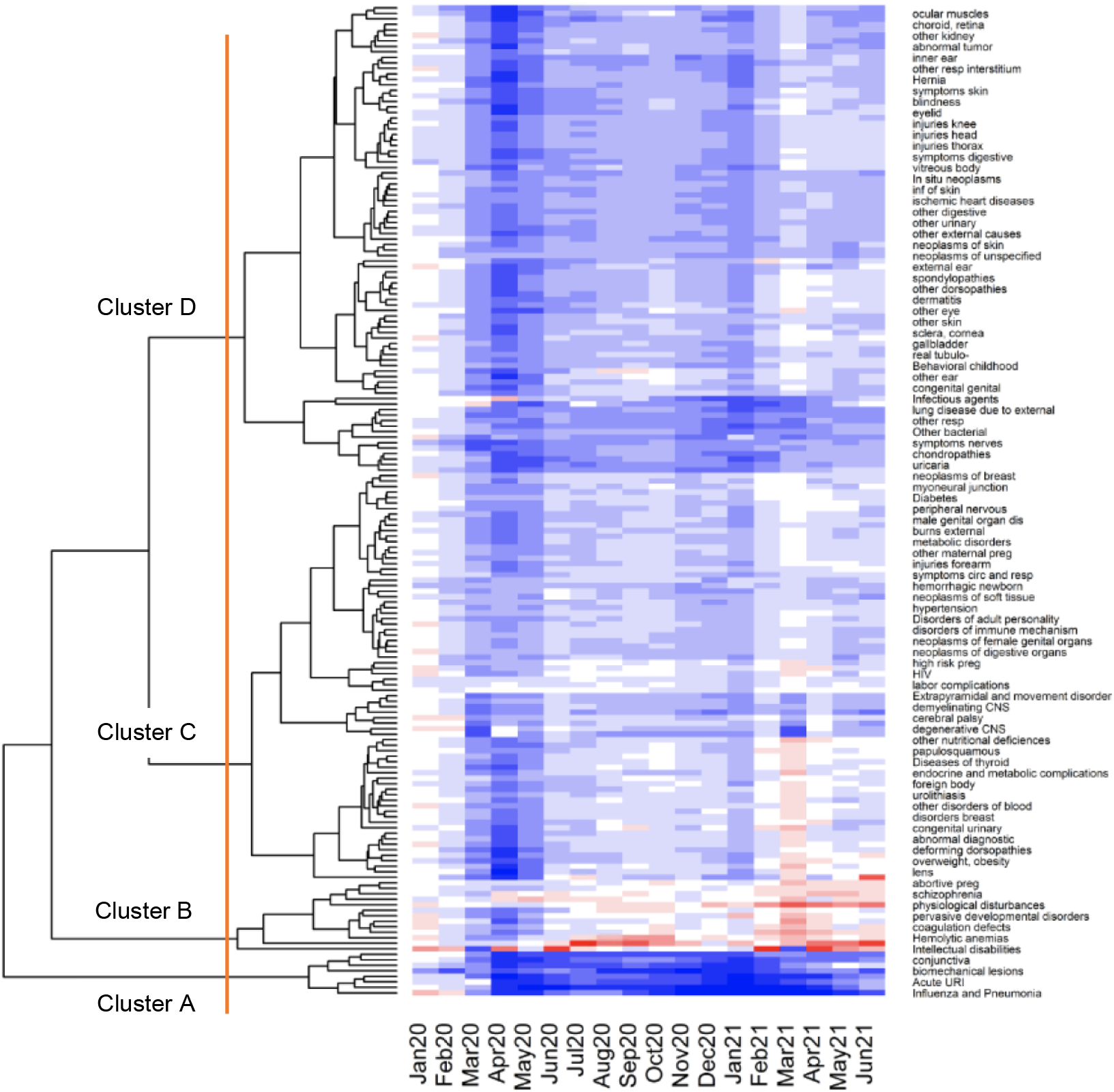
shows the clusters of the subchapters and where I made the cut-offs – can update this to have clear demarcations of the cut-offs

**Supplemental Figure S4.**
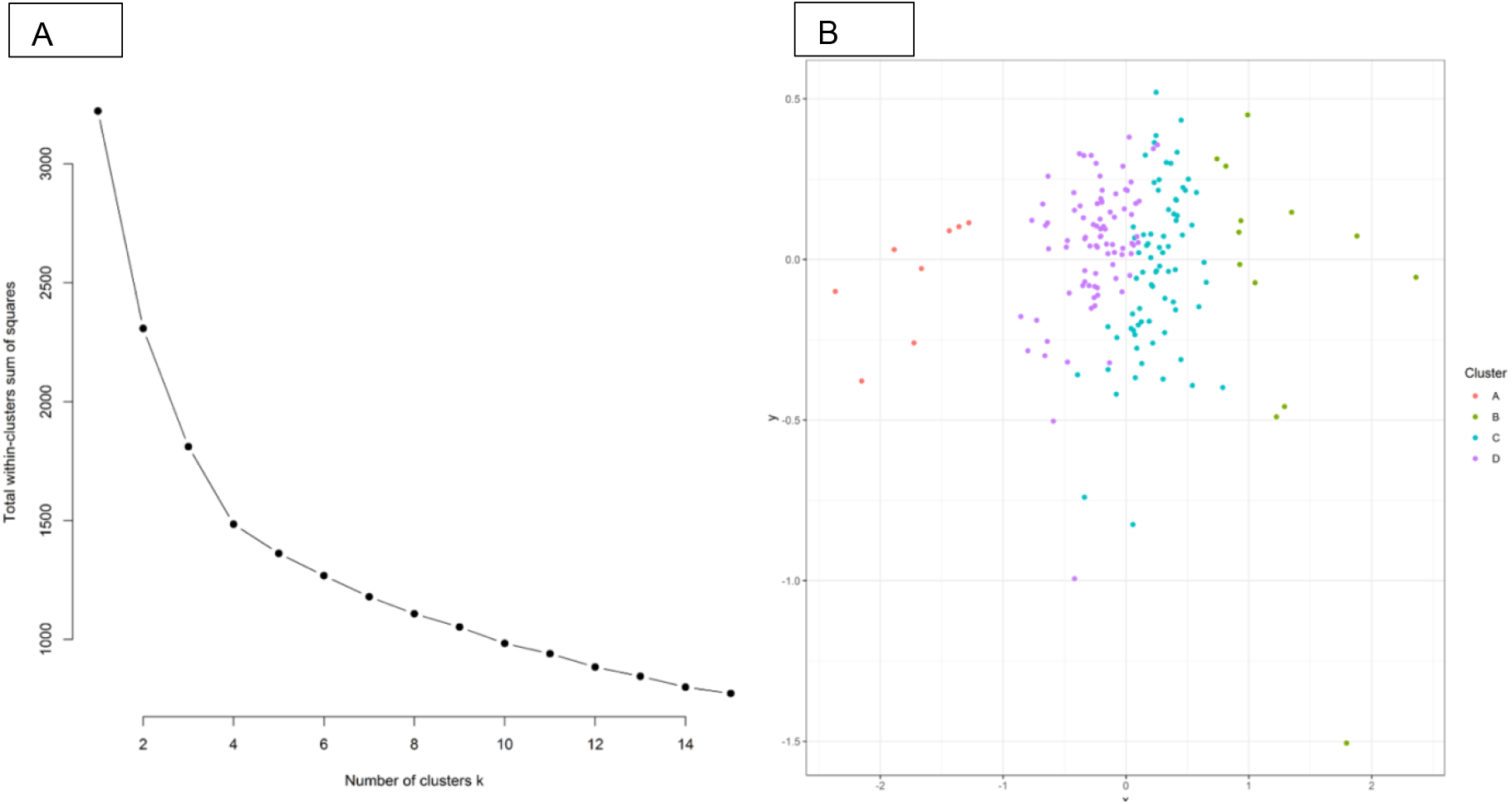
Results from k-means and principle components analysis of the ICD-10 subchapters Panel A shows an “elbow plot” with the explained within-cluster sum of squares on the Y-axis and the number of clusters on the X-axis. Panel B shows the principal components analysis of subchapters when categorized into 4 clusters. A cut-off of 4 clusters was chosen based on the elbow plot in panel A.

**Supplementary Table S4.** Incidence Rate Ratio and 95% Prediction interval of select Subchapters of disease, March 2021 to December 2021

| Subchapter | Mar | Apr | May | Jun | Jul | Aug | Sept | Oct | Nov | Dec |
| --- | --- | --- | --- | --- | --- | --- | --- | --- | --- | --- |
| I&P<br>(J09-J18) | 0.74<br>(0.72, 0.76) | 0.29<br>(0.28, 0.30) | 0.37<br>(0.35, 0.38) | 0.58<br>(0.56, 0.61) | 0.74<br>(0.71, 0.78) | 0.68<br>(0.65, 0.71) | 0.51<br>(0.49, 0.53) | 0.35<br>(0.34, 0.36) | 0.23<br>(0.23, 0.24) | 0.17<br>(0.17, 0.18) |
| Acute URI<br>(J00 – J06) | 1.09<br>(1.07, 1.08) | 0.33<br>(0.33, 0.34) | 0.28<br>(0.27, 0.28) | 0.39<br>(0.38, 0.40) | 0.58<br>(0.53, 0.59) | 0.51<br>(0.50, 0.52) | 0.51<br>(0.50, 0.52) | 0.52<br>(0.51, 0.53) | 0.46<br>(0.45, 0.46) | 0.35<br>(0.34, 0.35) |
| Acute LRI<br>(J20-J22) | 0.91<br>(0.87, 0.95) | 0.26<br>(0.25, 0.27) | 0.20<br>(0.19, 0.21) | 0.24<br>(0.22, 0.25) | 0.33<br>(0.31, 0.35) | 0.31<br>(0.29, 0.33) | 0.29<br>(0.27, 0.31) | 0.31<br>(0.30, 0.33) | 0.24<br>(0.23, 0.25) | 0.16<br>(0.15, 0.16) |
| Viral skin and<br>mucous<br>(B00 – B09) | 0.74<br>(0.69, 0.79) | 0.34<br>(0.32, 0.36) | 0.42<br>(0.40, 0.45) | 0.43<br>(0.41, 0.46) | 0.39<br>(0.37, 0.41) | 0.43<br>(0.40, 0.45) | 0.45<br>(0.43, 0.48) | 0.50<br>(0.47, 0.53) | 0.52<br>(0.49, 0.55) | 0.52<br>(0.49, 0.56) |
| Middle ear<br>(H65 – H75) | 0.66<br>(0.64, 0.68) | 0.19<br>(0.18, 0.19) | 0.30<br>(0.29, 0.31) | 0.41<br>(0.40, 0.43) | 0.53<br>(0.51, 0.55) | 0.59<br>(0.57, 0.61) | 0.59<br>(0.57, 0.61) | 0.50<br>(0.48, 0.52) | 0.40<br>(0.39, 0.42) | 0.32<br>(0.31, 0.33) |
| Intestinal<br>Infectious<br>(A00-A09) | 0.68<br>(0.64, 0.73) | 0.35<br>(0.32, 0.37) | 0.42<br>(0.39, 0.44) | 0.46<br>(0.43, 0.50) | 0.52<br>(0.48, 0.56) | 0.52<br>(0.49, 0.56) | 0.56<br>(0.53, 0.61) | 0.50<br>(0.47, 0.54) | 0.44<br>(0.41, 0.47) | 0.37<br>(0.34, 0.40) |
| Intellectual<br>Disabilities<br>(F70 – F79) | 0.51<br>(0.43, 0.60) | 1.77<br>(1.50, 2.11) | 1.04<br>(0.89, 1.24) | 2.04<br>(1.75, 2.42) | 1.30<br>(1.11, 1.53) | 1.21<br>(1.04, 1.43) | 1.08<br>(0.93, 1.27) | 1.10<br>(0.94, 1.29) | 1.20<br>(1.03, 1.44) | 1.15<br>(0.98, 1.38) |
| Schizophrenia<br>(F20 – F29) | 0.86<br>(0.82, 0.90) | 1.23<br>(1.17, 1.29) | 1.17<br>(1.12, 1.23) | 1.23<br>(1.17, 1.28) | 1.18<br>(1.13, 1.24) | 1.19<br>(1.14, 1.25) | 1.27<br>(1.21, 1.33) | 1.22<br>(1.16, 1.28) | 1.07<br>(1.02, 1.13) | 1.11<br>(1.06, 1.17) |
| STI<br>(A50 – A64) | 0.80<br>(0.71, 0.92) | 0.60<br>(0.53, 0.68) | 0.74<br>(0.66, 0.84) | 0.93<br>(0.83, 1.06) | 1.10<br>(0.97, 1.24) | 1.30<br>(1.16, 1.47) | 1.25<br>(1.11, 1.41) | 1.34<br>(1.20, 1.52) | 1.13<br>(1.01, 1.29) | 1.21<br>(1.07, 1.38) |
| Hemolytic<br>anemias<br>(D55 – D59) | 0.75<br>(0.67, 0.85) | 0.77<br>(0.68, 0.87) | 0.99<br>(0.88, 1.12) | 1.03<br>(0.91, 1.17) | 1.26<br>(1.12, 1.43) | 1.39<br>(1.24, 1.58) | 1.52<br>(1.35, 1.73) | 1.50<br>(1.33, 1.72) | 1.22<br>(1.09, 1.40) | 1.23<br>(1.09, 1.41) |
| Coagulation<br>defects<br>(D65 – D69) | 0.93<br>(0.83, 1.04) | 0.76<br>(0.68, 0.85) | 0.87<br>(0.78, 0.98) | 1.13<br>(1.02, 1.27) | 1.05<br>(0.95, 1.18) | 1.09<br>(0.98, 1.22) | 1.19<br>(1.07, 1.34) | 1.26<br>(1.13, 1.41) | 1.06<br>(0.95, 1.19) | 1.15<br>(1.02, 1.29) |

